# Characterising infectious disease mortality in severe mental illness: A retrospective matched cohort study

**DOI:** 10.1101/2025.10.08.25337061

**Authors:** A Ronaldson, J Das-Munshi, A Dregan, T Lampejo, C Henderson, Debs Smith, I Bakolis

## Abstract

**Background:** Evidence suggests that people with severe mental illness (SMI) are at an increased risk of infection mortality compared to the general population. Little is known about how this risk might differ across infection types, and the potential impact of sociodemographic and clinical factors. We investigated associations between SMI and infection mortality in a population-based cohort, examining variation by infection type and potential moderating factors.

**Study design:** This retrospective matched cohort study used national primary care data from the Clinical Practice Research Datalink (CPRD) from 1 January 2000 to 31 December 2019 linked with Office of National Statistics (ONS) mortality data. Competing risks regression and cause-specific hazard models assessed risk of infection mortality in people with SMI versus non-SMI controls. We examined risk across different infection types and assessed the impact of sociodemographic and clinical factors.

**Study results:** Our cohort comprised 84,494 people with SMI matched on age, gender, and GP practice with 84,494 non-SMI controls. Fully adjusted models showed that people with SMI were more likely to die from any infection compared to non-SMI controls (adjusted hazards ratio (aHR)=1.58, 95% confidence intervals (CI)=1.44 to 1.74). Infection-specific analyses revealed increased risk of death from respiratory (aHR=1.69, 95% CI=1.51 to 1.89), gastrointestinal (aHR=2.01, 95% CI=1.16 to 3.48), and renal/urinary (aHR=1.70, 95% CI=1.32 to 2.19) infections in the SMI group.

**Conclusions:** People with SMI are at increased risk of infection mortality, especially from respiratory, gastrointestinal, and renal/urinary infections. We recommend prioritising this group for preventative measures including influenza and pneumococcal vaccines.

## Introduction

Evidence suggests that people with SMI may also be at an increased risk of death from infectious diseases, when compared with the general population. Early in the COVID-19 pandemic, it emerged that people with SMI were more likely to die from COVID than those without SMI, which has since been confirmed by numerous reviews and meta-analyses^1–8^. In recent years, a number of reviews have emerged showing that people with SMI are at increased risk of dying from infectious diseases more generally^9–11^, and that this mortality risk is considerably pronounced for respiratory infections, particularly pneumonia^11^. Results have been more mixed for sepsis mortality^11,12^, which warrants further investigation. Several sociodemographic and clinical factors may moderate risk of death from infection in this population. For example, some studies reported that infection mortality risk is more pronounced in men with SMI^11^, and there is a consistent association between deprivation and poor infection outcomes^13^. Studies have reported that COVID-19 mortality risk in SMI was moderated by ethnicity with risk highest among people from Black ethnic groups when compared to people from White ethnic groups^14,15^. However, ethnic differences in risk of dying from other infections in people with SMI beyond COVID-19 remain underexplored.

Additionally, there is also evidence suggesting that the risk of infection mortality might differ across specific SMI diagnoses^16,17^.

### Study aims

Therefore, this study aimed to comprehensively examine the association between SMI and infectious disease mortality within a population-based cohort. We investigated whether this association varied by type of infection and assessed potential effect modification by gender, deprivation levels, ethnic background, and specific SMI diagnoses.

## Methods

### Study design and population

We implemented a retrospective matched cohort study design using data from the Clinical Practice Research Datalink (CPRD) Aurum database. The CPRD Aurum database is one of the largest electronic primary care databases globally, containing historical data from general practices in England and Northern Ireland for a considerable subset of the UK population (approximately 24%). It has been shown to be broadly representative of the UK population regarding age, gender, area-level deprivation, and geographical distribution^18^. The CPRD contains detailed information on clinical diagnoses, therapies, referrals, laboratory tests, and sociodemographic variables, and established data linkages with Office of National Statistics (ONS) mortality data (England only) and Hospital Episode Statistics (HES). Ethical approval for this study was granted by the CPRD Independent Scientific Advisory Committee (protocol no. 22_001763).

The study period ran from 1 January 2000 to 31 December 2019. This study end date was chosen to avoid the potentially confounding effects of the COVID-19 pandemic on other infection mortality. We included patients with a first diagnostic record of SMI in the study period (incidence cohort) using medical codes for schizophrenia, schizoaffective disorders, bipolar disorder, and other psychoses. Patients entered the cohort on the date of first SMI diagnostic record (study index date) and had to be 16 years or older at this time. Matched control patients were assigned the same index date as their corresponding SMI patient.

Patients exited the cohort at date of death, or 31 December 2019, whichever occurred first. Patients 100 years or older at SMI diagnosis were excluded from the study. Only patients with linkage available to ONS mortality data were included in the study, leading to the inclusion of English GP practices only. Patients with SMI were matched 1:1 on age at index date for SMI, gender, and primary care practice with control patients without SMI. This study is reported as per the Strengthening the Reporting of Observational Studies in Epidemiology (STROBE) guidelines.

### Outcome: Infectious disease mortality

Detailed information about cause of death were made available through linkage with ONS death registration data. The ONS provides data for the causes of death and the exact date of death as recorded on death certificates by registered medical practitioners. Death certificates in England record primary cause of death as well as any secondary or contributing conditions (typically three but can be more) that may have led to the primary cause. Causes and conditions are recorded using International Classification of Diseases, Tenth Revision (ICD-10) codes.

The main outcome in the current study was death from any infection as a primary cause of death. We also investigated death from specific infection types which included respiratory (e.g. pneumonia, influenza), sepsis, gastrointestinal, renal and urogenital, central nervous system (CNS), HIV or hepatitis, skin, and other infections (e.g. infectious arthropathies).

Lists of ICD-10 codes used to define outcomes are provided in Supplementary Material Table S1.

### Covariates

Variables associated with the risk of SMI as well as infection mortality in the literature were chosen for inclusion as covariates. These variables included (in addition to matched variables) ethnicity, area-level deprivation, body mass index (BMI), smoking status, and the presence of long-term conditions. Ethnicity was obtained from primary care data in the first instance. If this information was missing in primary care, it was obtained from linkage with HES where possible. Ethnicity was categorised into Asian (Bangladeshi, Indian, Pakistani, and other Asian), Black (African, Caribbean, Other), Mixed, Other (e.g. Middle Eastern, Central American), and White. Area-level deprivation was measured using the English Index of Multiple Deprivation (IMD) at the lower-layer super output area (LSOA) level. IMD provides a metric of deprivation based on seven domains using specific weightings: education, employment, income, health, barriers to housing and services, living environment, and crime^19^. We created IMD quintiles (1=least deprived, 5=most deprived). Smoking status was based on the status recorded closest to the study index date (i.e. when SMI diagnosis was first recorded). Patients were categorised as current smokers, ex-smokers, or never smoked. BMI (kg/m^2^) was based on the record made closest to study index date. Patients were categorised as underweight (<18.5 kg/m^2^), normal weight (18.5 to 24.9 kg/m^2^), overweight (25 to 30 kg/m^2^), or obese (>30 kg/m^2^). Long-term physical and mental health conditions recorded in primary care records before the patient’s study end date were also included as a covariate. We included conditions linked to an increased risk of poor infection outcomes: anxiety, asthma, atrial fibrillation, autoimmune disorders (colitis, Crohn’s disease, lupus, rheumatoid arthritis, psoriasis), cancer, chronic obstructive pulmonary disorder (COPD), coronary heart disease (CHD: angina, myocardial infarction, heart failure), depression, dementia, eating disorder, epilepsy, gastroesophageal reflux disorders, hypertension, ischaemic stroke, kidney disease, liver disease, and substance use disorders (both alcohol and drug), type 2 diabetes mellitus^15^.

We initially proposed to examine differential risk across genders, ethnic groups, IMD quintiles, and SMI diagnoses *a priori*. SMI patients were grouped into the following diagnostic groups: ‘Schizophrenia/psychosis’ (including schizophrenia, schizoaffective disorder, and/or other psychoses), ‘Bipolar disorder’, and ‘Both diagnoses’ (patients with primary care records of both schizophrenia/psychosis and bipolar disorder). Consultation with the study’s LEAP highlighted the potential role of antipsychotics and antidepressants in infection mortality risk among SMI patients. In response, we also examined rates of antipsychotic and antidepressant prescribing throughout each patient’s study period (first SMI record to study end date) and assessed differential risk across those who had and had not received a prescription among the SMI group only – this was considered *a posteriori*.

A full list of codes for covariates is available on request from the authors.

### Statistical analysis

We used descriptive statistics to summarise differences in sample characteristics and mortality between SMI cases and matched controls.

We first assessed unadjusted associations in the matched cohort, followed by fully adjusted models incorporating the complete set of covariates. For unadjusted models, we used competing risks regression models to estimate differences in infection mortality between patients with and without SMI. Mortality from other causes was treated as a competing risk. When looking at mortality from specific infection types (e.g. respiratory infections), mortality from other infections as well as non-infection causes was the competing risk.

For fully adjusted analyses, data were missing for several covariates: BMI (28.5%), ethnicity (15.6%), IMD (0.3%), and smoking status (18.9%). Multiple imputation using chained equations with five imputations was performed to handle missing data. The imputation model included all covariates as well as predictors (SMI diagnosis) and outcomes (death). It was not possible to perform fully adjusted competing risks regression analyses on imputed datasets due to the computational power required. Therefore, we applied cause-specific hazard models to imputed data to estimate differences in infection mortality between patients with and without SMI, while accounting for mortality from non-infectious causes^20^. The proportional hazards assumption was assessed by plotting Schoenfeld residuals using unimputed data^21^.

To assess whether risk of death from any infection in those with SMI differed across gender, ethnicity, neighbourhood deprivation, and SMI diagnosis, we included interaction terms (e.g. SMI*gender) in fully adjusted cause-specific hazard models. When significant interactions were identified, we conducted stratified analyses to explore these differences further. To assess the impact of the prescription of antipsychotics and antidepressants on risk of death from any infection we performed fully adjusted (including age and gender) cause-specific hazard models in the SMI patient group only.

All analyses were conducted using STATA 18.0 (Stata Corp LLP, College Station, Texas, US).

### Sensitivity analysis

For a broader assessment of infectious disease mortality, we repeated all analyses with the outcome being death with infection as the primary or secondary/contributing cause of death.

To ensure more long-term assessment of the impact of SMI on infection mortality, we repeated fully adjusted analyses excluding those with less than one year of follow-up.

As the missing-at-random assumption might not hold when using routinely collected data, we also carried out complete-case analysis to confirm findings from imputed estimates. These analyses were fully adjusted and also adjusted for age and gender as removing all participants with missing data led to case-control imbalance in terms of matching.

In fully adjusted models, Schoenfeld residuals indicated that proportional hazard assumptions had been violated for some covariates. This is not surprising as testing this assumption relies on failure to reject the null hypothesis which becomes less likely with larger sample sizes. Therefore, we performed a post-hoc sensitivity analysis using logistic regression to model fully adjusted associations between SMI status and risk of death from infections, where time-to-event was not considered. This approach allowed us to verify the robustness of our findings, although it did not account for the timing of events.

### Involvement of people with lived experience

This study was supported by a Lived Experience Advisory Panel (LEAP) of five people with relevant lived experience. They met four times with the first author (AR) to plan and discuss the study. LEAP members contributed to shaping the research question, guiding the analysis, interpreting results, and drafting the final write-up.

### Role of the funding source

This work was supported by MQ Mental Health Research Fellowship (MGF22\12). JDM and AD are partly funded by the UKRI MRC PROMISE consortium. AD is partially funded by the National Institute of Health and Care Research (NIHR) (grant: NIHR203988). The views expressed are those of the authors. For the purposes of open access, the author has applied a Creative Commons Attribution (CC BY) licence to any Accepted Author Manuscript version arising from this submission.

## Results

### Sample selection and characteristics

The final study sample comprised 84,494 patients with SMI, and 84,494 non-SMI controls matched on age, gender, and GP practice. The selection of patients into the study is presented in Figure 1. The maximum study follow-up period was 20 years, with an average of eight years (SD=5.7 years) follow-up. Sociodemographic and clinical sample characteristics are presented in Table 1. A higher proportion of patients with SMI were of Black ethnicity and were from areas with higher deprivation, compared with the non-SMI control group. More patients with SMI were overweight and obese, were smokers, and had multiple LTCs than control patients. Most patients with SMI had a primary care record of schizophrenia/psychosis (66.5%), 27.2% had a record of bipolar disorder, and 6.3% had a record of both. The rate of all-cause mortality over the study period was higher in people with SMI than people without SMI (SMI: 19.1%, non-SMI: 10.8%).

**Figure 1.**
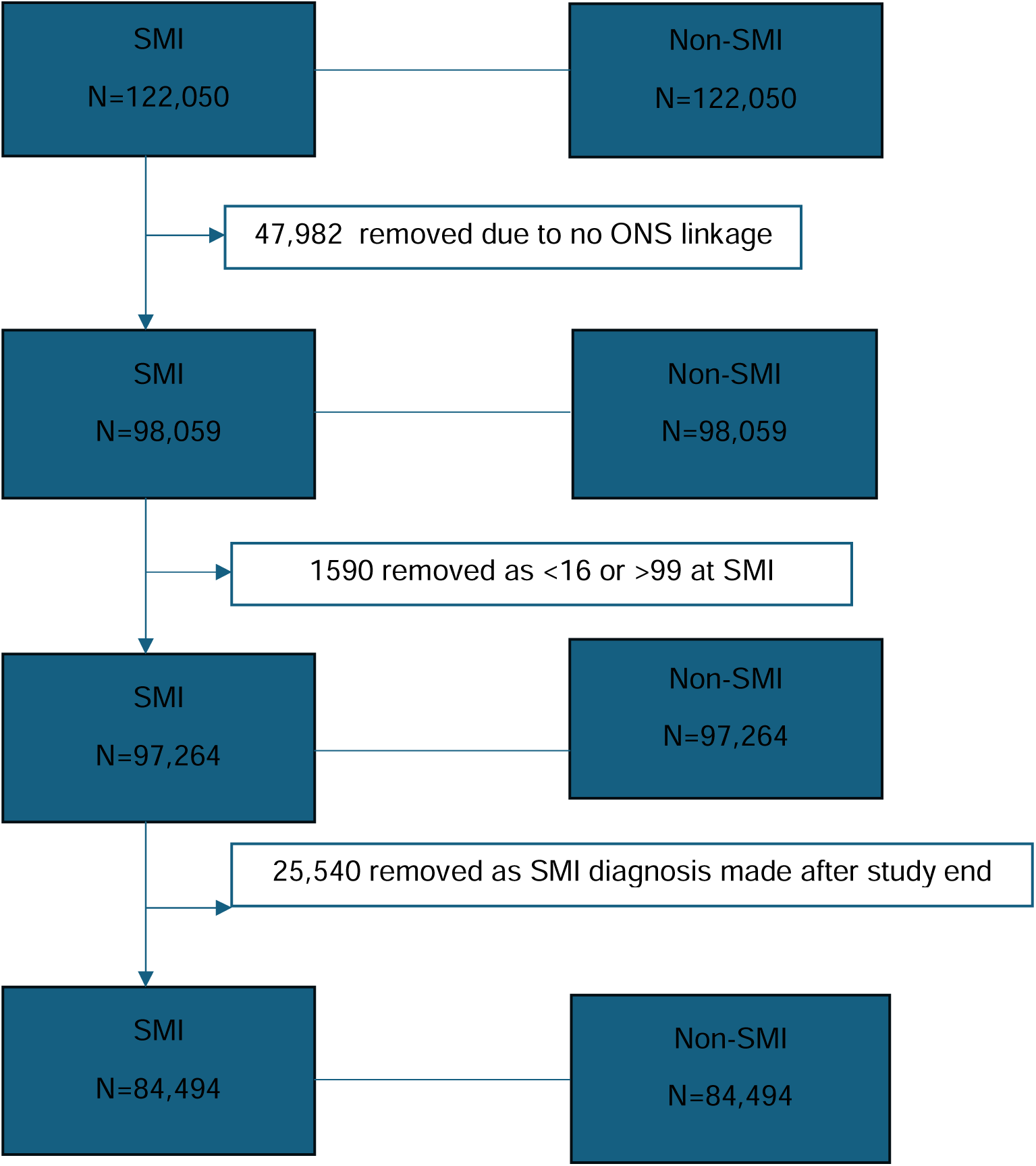
Flow diagram illustrating the selection of patients into the study.

**Table 1.**
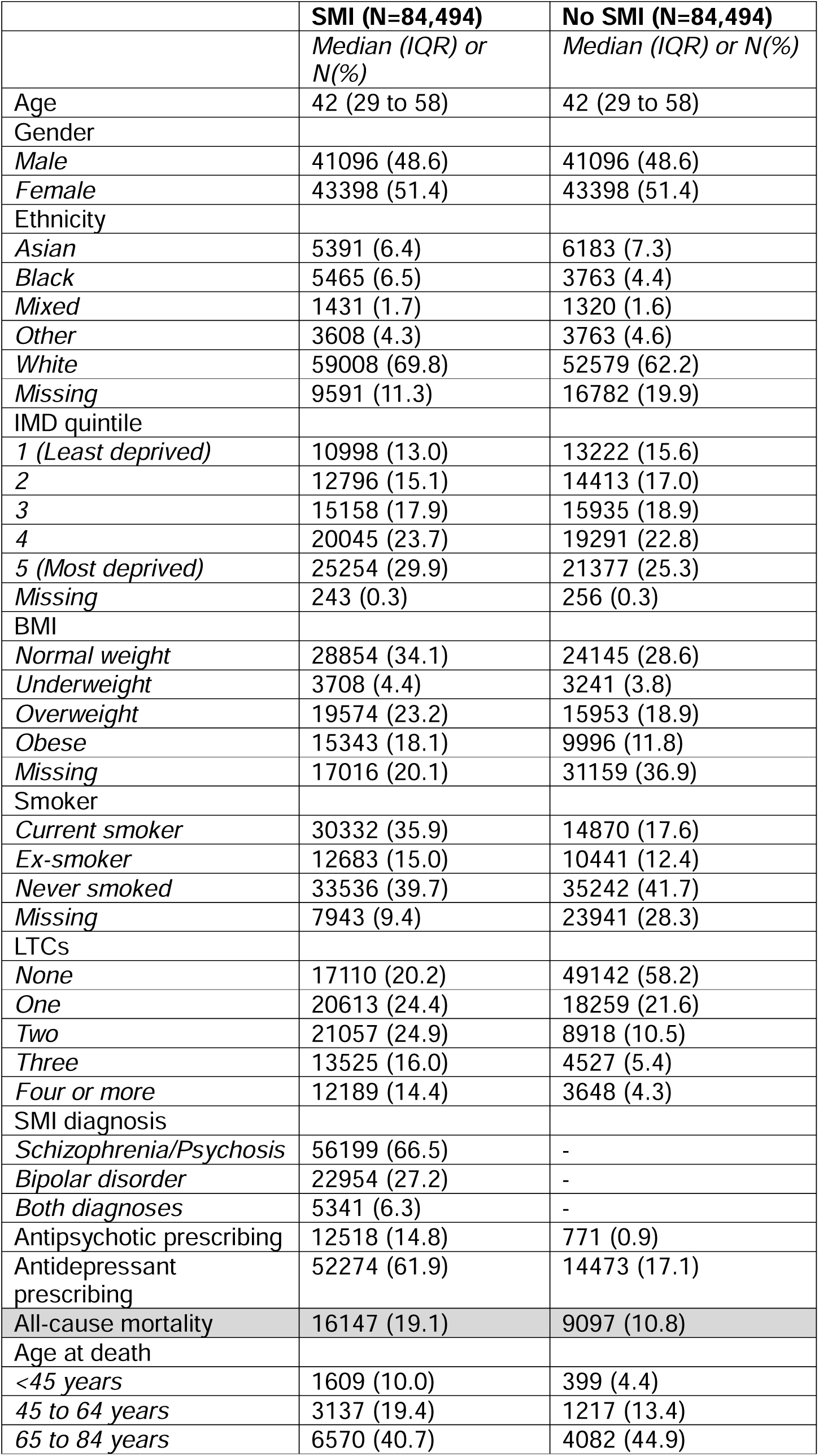
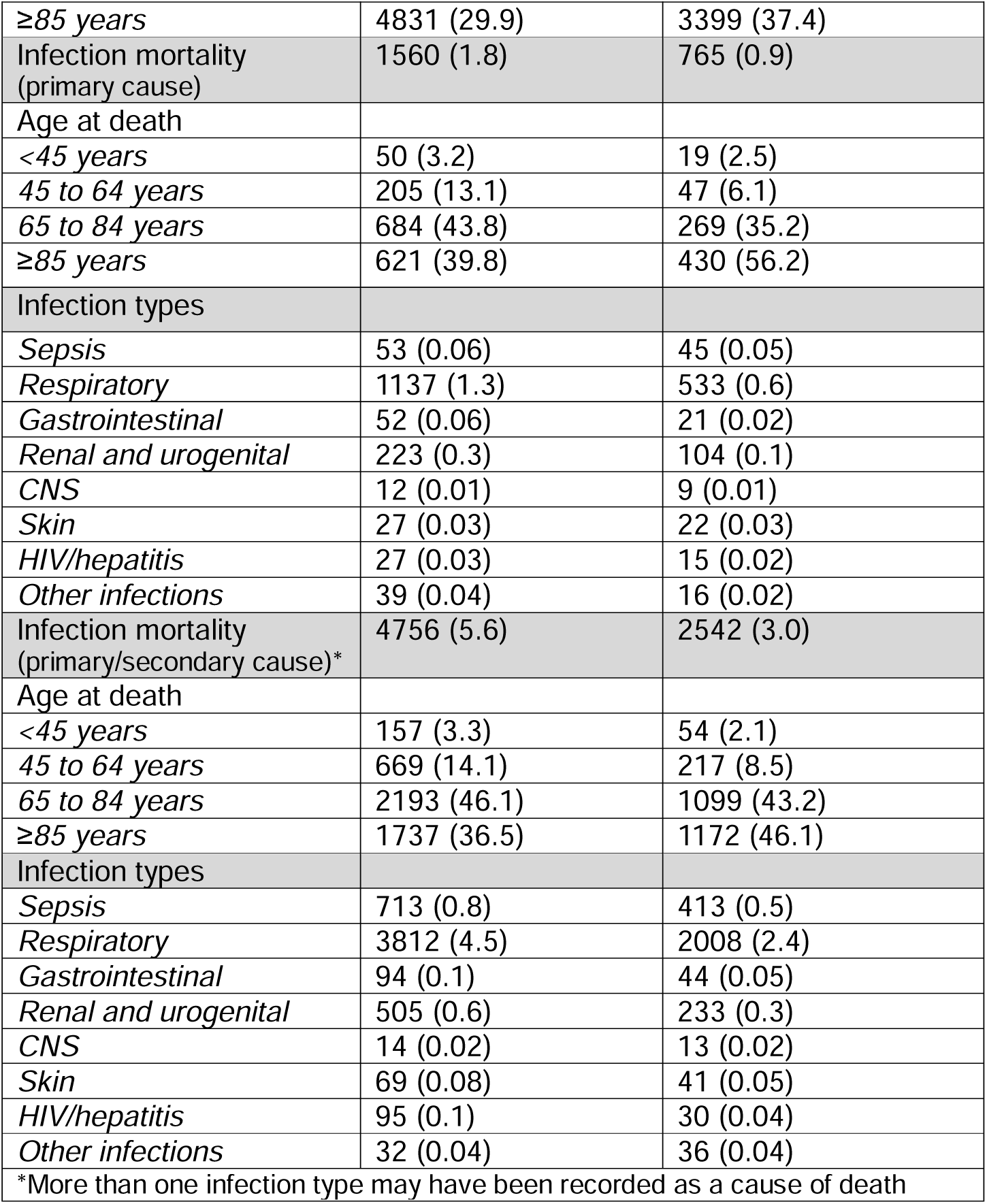
Sample characteristics.

Infection mortality rates are presented in Table 1. Double the number of patients with SMI died from any infection compared to non-SMI controls (SMI: 1.8%, non-SMI: 0.9%), and this difference was most pronounced amongst those who died in the middle (45 to 64 years) to older (65 to 84 years) age group. In terms of mortality rates for specific infection types, a higher proportion of patients with SMI died from all infection types compared with non-SMI controls, except for CNS and skin infections. 14.8% of patients with an SMI record had received an antipsychotic prescription within primary care, and 61.9% had received an antidepressant prescription, since their SMI diagnosis.

### Risk of infectious disease mortality in people with SMI

Unadjusted and fully adjusted hazard ratios for mortality from infection as a primary cause of death are presented in Figure 2. Fully adjusted models revealed that people with SMI were 58% more likely than non-SMI controls to die from any infection (aHR=1.58, 95% CI=1.44 to 1.74). Looking at infection subtypes showed that those with SMI were at increased risk of death from respiratory infections (aHR=1.69, 95% CI=1.51 to 1.89), gastrointestinal infections (aHR=2.01, 95% CI=1.16 to 3.48), and renal/urinary infections (aHR=1.70, 95% CI-1.32 to 2.19). There was no significantly increased risk from the other infection types (sepsis, CNS, skin, HIV/hepatitis, other infections).

**Figure 2.**
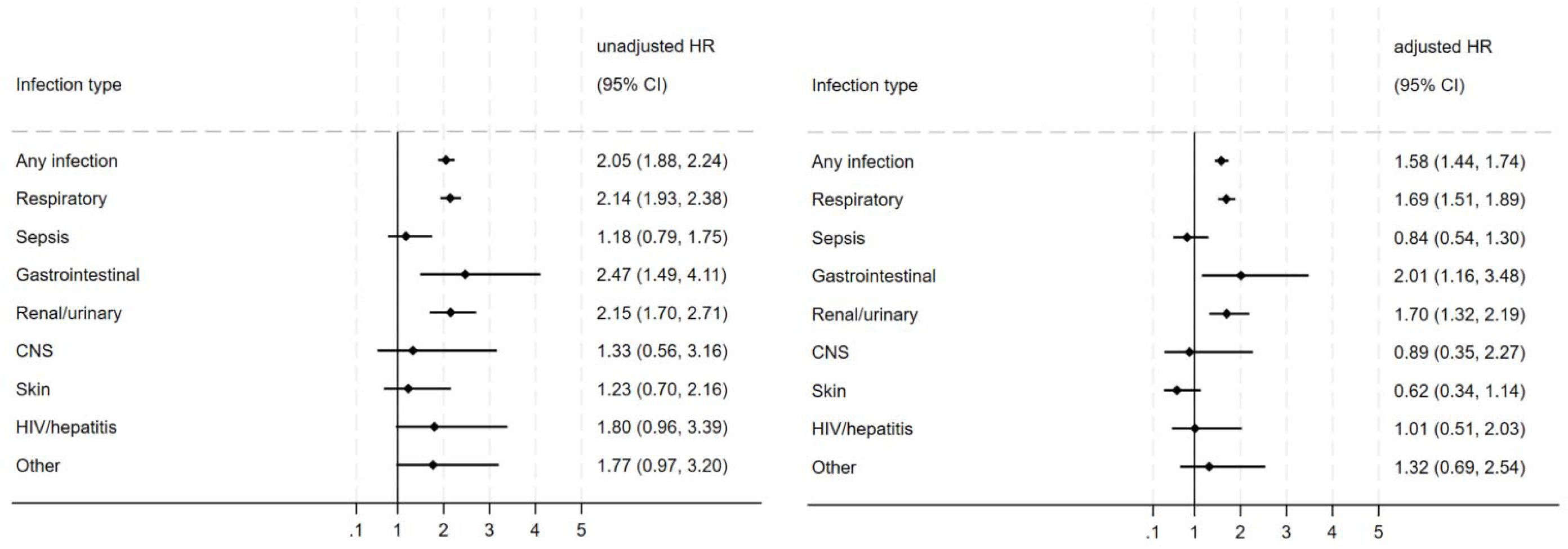
Hazard ratios for the association of severe mental illness (SMI) with infection mortality overall and mortality from infection subtypes (as primary cause of death). Unadjusted estimates are from competing risks regression models on unimputed data. Adjusted estimates are from cause specific hazard models on imputed data. Unadjusted models were based on an age-, sex-and GP practice-matched cohort. Fully adjusted models were further adjusted for ethnicity, neighbourhood deprivation, BMI, smoking status, and number of long-term conditions.

Fully adjusted analyses revealed significant interactions between SMI and IMD, and SMI diagnosis, (Table S2 in Supplementary Material). Stratified analyses are presented in Figure 3. Results indicate that risk of death from any infection was highest among people with SMI from the least deprived areas (aHR=2.10, 95% CI=1.65 to 2.64), although risk among people with SMI compared to non-SMI controls was increased across all IMD quintiles. Stratifying by SMI diagnosis revealed that risk of infection mortality was significantly increased in patients with a record of schizophrenia/psychosis (aHR=1.99, 95% CI=1.78 to 2.22). Patients with bipolar disorder were at no increased risk of dying from infection and risk was significantly lower in patients with both diagnoses compared to controls (aHR=0.66, 95% CI=0.45 to 0.97). In SMI patients only, we found that receipt of a prescription for an antipsychotic was associated with increased risk of dying from any infection (aHR=1.48, 95% CI=1.32 to 1.65), but antidepressant prescribing had no significant impact (p=0.198).

**Figure 3.**
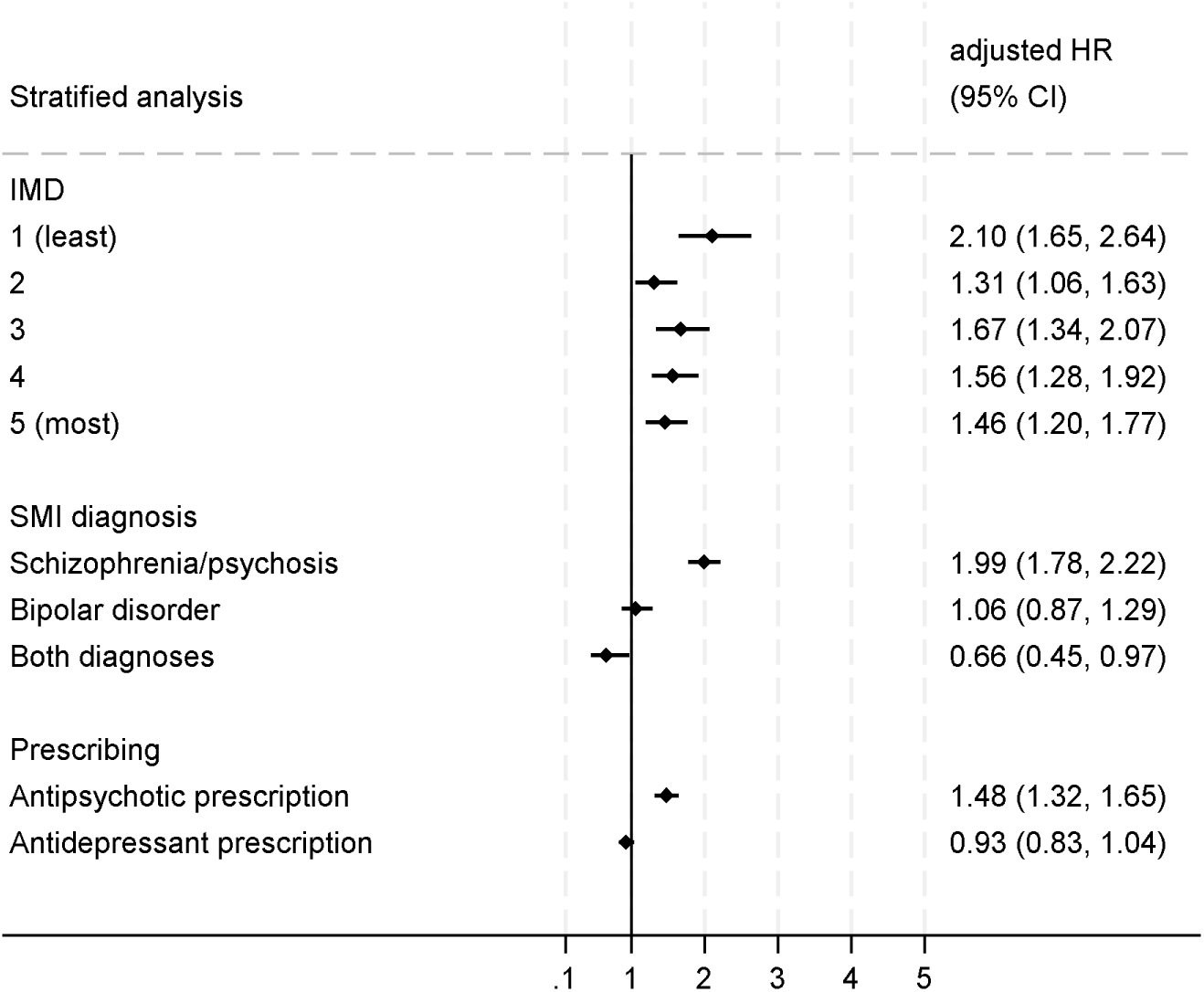
Stratified analysis. Hazard ratios for the association of severe mental illness (SMI) with infection mortality (any infection) across IMD quintiles and SMI diagnosis. For these stratified analyses, patients without SMI are the reference group. The impact of antipsychotic and antidepressant prescribing on infection mortality is assessed in SMI patients only. For these analyses, SMI patients who have no prescription record for the drug type of interest serve as the reference group. Estimates are from fully adjusted cause specific hazard models on imputed data.

### Sensitivity analysis

Infection mortality rates (primary and/or secondary cause of death) are presented in Table 2. Unadjusted and fully adjusted hazard ratios for rates of mortality from infection as a primary or secondary/contributing cause of death are displayed in Figure S1 (Supplementary Material). Patients with SMI were 33% more likely than non-SMI controls to have an infection recorded as a primary or secondary cause of death (aHR=1.33, 95% CI=1.26 to 1.40). Risk was increased for death from respiratory, gastrointestinal, and renal/urinary infection mortality, as well as sepsis-related death.

We repeated fully adjusted analyses excluding patients with less than one year of follow-up (Figure S2). People with SMI were 63% more likely than non-SMI controls to die from any infection (aHR=1.63, 95% CI=1.46 to 1.81). Looking at infection subtypes showed that those at SMI were at increased risk of death from respiratory infections (aHR=1.72, 95% CI=1.52 to 1.96), and renal/urinary infections (aHR=1.87, 95% CI=1.40 to 2.48). There was no significantly increased risk of death from the other infection types.

We performed a complete case analysis to confirm findings from imputed estimates. Results are presented in Figure S3. Complete case estimates more pronounced than imputed estimates but follow the same pattern.

Results of logistic regressions to verify the robustness of our findings in the face of proportional hazard assumption violation are presented in Figure S4 and were similar to those produced using cause-specific regression hazard models.

## Discussion

Using data from a nationally representative matched cohort we found that people with SMI had a 58% higher risk of dying from any infection compared to those without SMI. Further examination of infection types revealed an elevated risk of death from respiratory, gastrointestinal and renal/urinary infections specifically. This discrepancy in mortality risk between those with and without SMI appeared to be most pronounced in middle-age (45-64 years). Examination of potential sociodemographic factors showed that infection mortality risk was highest among individuals with SMI living in the least deprived areas, but gender and ethnicity did not modify infection mortality risk. When analyses were stratified, we found that risk of death from infection was increased for people with schizophrenia and/or psychosis, but not for people who had a primary care record of bipolar disorder. When we looked at the SMI patient group only, we found that people who had received a prescription for antipsychotics were at an increased risk of death from any infection, but antidepressant prescription inferred no increased risk.

The main finding of the current study is in line with previous meta-analyses which have reported increased risk of infection mortality in SMI ^9–11^. However, effect sizes reported in this study are lower than pooled effects reported previously. This discrepancy may be partly due to previous studies predominantly defining SMI cohorts based on data from secondary mental health services. In contrast, the current study utilizes primary care data, potentially capturing a more comprehensive cohort that includes individuals both engaged with and outside of secondary services. Findings from these meta-analyses also align with the results of this study relating to respiratory infection mortality risk in SMI, but again, effect sizes reported in this study are lower than previously reported pooled effects^9,11^. The two-fold increase in gastrointestinal infection mortality risk reported in the current study conflicts with those reported using national data from Wales which found no increased risk of intestinal infections in people with SMI^22^. Despite wide confidence intervals, the consistency of the observed effect suggests a genuine signal of risk, even of the precise magnitude remains uncertain. To the best of our knowledge, we are the first to report increased renal/urinary infection mortality in people with SMI, although our finding does align with research showing higher prevalence of chronic kidney disease (a major driver of renal and urinary infections) in this population^23^. Moreover, there is evidenced for increased risk of urinary infection in acute psychosis^24^.

We found that individuals with SMI were not at increased risk of sepsis mortality as a primary cause of death but had a higher risk when sepsis was listed as a secondary or contributing factor. This mixed result aligns with previous meta-analyses which have reported decreased, unchanged, or increased sepsis mortality risk in SMI, depending on study design^11,12^. A recent genome wide association (GWAS) study found no significant causal evidence linking SMI to sepsis mortality, though there were indications of genetic associations with sepsis incidence^25^. These findings suggest that sepsis may serve as a proxy for broader immune dysfunction in SMI contributing to increased susceptibility while not necessarily elevating mortality risk. Further research is needed to understand sepsis incidence and outcomes in this population.

Despite adjusting for several relevant sociodemographic and clinical factors, risk of infection mortality remained elevated in people with SMI suggesting that additional mechanisms may contribute to this increased risk. One such mechanism relates to the significant inequalities people with SMI experience around healthcare service provision and stigma within healthcare settings. For example, people with SMI can experience ‘diagnostic overshadowing’ where presenting symptoms of a physical illness are judged to be a manifestation of the mental illness^26^. Experiences of diagnostic overshadowing as well as experiences of stigma in healthcare settings might dissuade help seeking and delay receipt of appropriate care^27^, which is known to be a risk factor for poor infection prognoses^28,29^.

SMI-related factors such as diagnosis and severity are likely relevant also. In the current study we found that risk of death from infection was increased in those with primary care records of schizophrenia and psychosis, but not in those with records of bipolar disorder, which is in line with previous research^16,17^. Although schizophrenia and bipolar disorder share certain symptoms, such as depression, negative symptoms are a distinguishing feature of schizophrenia and often persist despite treatment^30^. Negative symptoms such as apathy and avolition might increase the risk of self-neglect and reduced healthcare-seeking behaviour, potentially worsening infection prognoses. In contrast, bipolar disorder typically follows a relapse-remission pattern, allowing individuals to experience periods of stability between episodes, which may positively influence health outcomes.

There are biological changes associated with having SMI that might contribute to increased infection mortality in this population. First, SMI is characterised by several systemic immunological and inflammatory changes which likely have implications for infection incidence and prognosis. For example, there is evidence that people with psychotic disorders experience broad activation of the immune system^31^, have increased levels of peripheral proinflammatory cytokines^32^, as well as abnormal lymphocyte subpopulation counts^33^. Furthermore, people with SMI show attenuated responses to vaccines providing further evidence for immunocompromise^34^. A second biological pathway which might contribute to increased infection mortality risk in SMI are changes to the gut microbiome.

There is evidence people with SMI have increased microbial disorganisation in the gut, increases in bacterial strains associated with gastrointestinal inflammation, alongside decreases in anti-inflammatory strains^35^. Gut microbiota are known to play a role in gastrointestinal infections^36^, which is particularly pertinent to the increased risk of gastrointestinal infection mortality we report in the current study.

We found that patients with SMI who had received an antipsychotic prescription had an increased risk of infection-related mortality. This contrasts with a recent meta-analysis, which reported lower all-cause mortality among people with schizophrenia using antipsychotics compared to non-users^37^. This discrepancy may suggest that antipsychotic usage confers a specific disadvantage in infection-related outcomes. For instance, clozapine has been linked to increased infection risk, particularly respiratory and gastrointestinal infections^38^.

Additionally, antipsychotic use is associated with higher pneumonia risk^39^ and pneumonia mortality^40^. Potential mechanisms include antipsychotic-induced immunomodulation and immunosuppression^41,42^, which may directly elevate infection risk. Further uncertainty around interactions between various antipsychotic medications and antimicrobial agents could contribute to prescribing hesitancy, delayed treatment, and consequently poor infection outcomes. Given these concerns, the impact of antipsychotics on infection-related mortality warrants further investigation.

We found that infection mortality risk was modified by area-level deprivation. Although risk was increased across all deprivation quintiles, it was most pronounced in people with SMI living in the least deprived areas. More affluent areas tend to have older populations, which could contribute to increased infection mortality; however, given that our study cohort was age-matched, other unmeasured factors might be relevant. It is possible that individuals from more deprived areas exhibit different patterns of underlying long-term conditions that exacerbate infection mortality risk. Further research is needed to clarify the role of specific health conditions in the relationship between SMI and infection mortality.

### Strengths and limitations

Strengths of this study include the prospective study design with follow-up of up to twenty years. As most of the general population in England is under primary care and this is available free at the point of use, our sample is likely representative of the population in terms of age, gender, and ethnicity suggesting good generalisability of results^43^. We believe the use of primary care data over secondary mental health care data in this instance is a strength of this study, allowing for a more comprehensive SMI cohort. Incidence of SMI in primary care samples has been found to be similar to incidence rates in the community^44^ meaning we are confident our sample is representative of the SMI population in England.

However, we note that some populations were likely not represented in our sample such as prisoners and people experiencing homelessness^43^. The breadth of primary care records allowed us to adjust for several relevant confounding factors and to explore the role of modifying factors, but we cannot exclude the possibility of residual confounding. Although linkage with ONS mortality data allowed for comprehensive assessment of infection mortality, we may have lacked sufficient power to identify associations between SMI status and mortality from sepsis, CNS infections, skin infections, HIV/hepatitis, and other infections, as these were rarely recorded as primary cause of death. When we expanded our analysis to include infection mortality as either primary or secondary, we observed a significant increase in sepsis-related mortality among SMI patients. This association then reached statistical significance, suggesting that the initial lack of detection may have been due to limited power.

## Conclusion

Our analysis of national primary care data linked to the national mortality register provides clear evidence that the health inequalities faced by individuals with SMI extend to infection-related mortality, with a particularly heightened risk for respiratory, gastrointestinal, and renal/urinary infections. Given these findings, we strongly recommend that people with SMI in England be prioritized for preventative measures such as influenza and pneumococcal vaccines. Moreover, strategies are needed to address the low pneumococcal and influenza vaccine rates and high vaccine hesitancy in adults with psychiatric disorders^45^.

### Lived Experience Commentary

As someone with lived experience of mental health issues the fact that we finally have robust research that those with severe mental health issues are likely to die and get very ill at a younger age than their peers who do not have severe mental issues is really important. I am on antipsychotic medication, and this led me to have a greater risk of developing diabetes, and I did indeed develop this, and it has had an impact on my overall wellbeing. I wish that this research had been done before and the fact that it has been done is good and it has the potential to positively impact the lives of many people with severe mental health issues.

However good and relevant this research is it cannot improve things without the help and co-operation of policy makers and indeed those who implement policy and those who educate clinicians. My recommendation is that policy makers institute education for all those who are diagnosed with severe mental health issues so that they know the greater risk that they have in comparison with those who do not have severe mental health issues of developing physical health issues and of dying younger. I would suggest they are also educated about healthy lifestyle choices and are given tailored support which should include access to dieticians, physiotherapists and psychologists to try and follow what they have learnt. I would suggest they have yearly physical health checks including blood tests. I would suggest much more education for clinicians, including the findings of this research, and how to spot the early signs of physical health issues.

Finally, I suggest that further research connect with the Advanced Discovery Pain Platform (https://apdp.community/) and its consortium which looks at chronic pain and its causes. Understanding how pain affects mental health, the role of specific comorbidities, and whether the medications patients take increase the risk of serious physical health issues, and even earlier death, will give us crucial insights and inform targeted education for clinicians. Finally, this would also mean that people have better understanding of the illnesses they might get and be more motivated to follow the education and tailored support I suggested earlier.

## Declaration of interests

None to declare

## Funding

This work was supported by MQ Mental Health Research Fellowship (MGF22\12). JDM and AD are partly funded by the UKRI MRC PROMISE consortium. AD is partially funded by the National Institute of Health and Care Research (NIHR) (grant: NIHR203988). The views expressed are those of the authors. For the purposes of open access, the author has applied a Creative Commons Attribution (CC BY) licence to any Accepted Author Manuscript version arising from this submission.

## Acknowledgments

We would like to express our gratitude to the members of the project’s Lived Experience Advisory Panel for their valuable insights and contributions to this paper.

## Author contributions

AR, AD, JD-M, TL, and IB conceived the study and developed the study design. AD curated the data and AR and AD led and performed all analyses. AR led the draft of the manuscript with a contribution from DS, and all authors contributed to the interpretation of findings. All authors read, commented, and approved the final version of the paper.

## Transparency declaration

The lead author (AR) and the manuscript guarantor (IB) affirm that the manuscript is an honest, accurate, and transparent account of the study being reported, no important aspects of the study have been omitted, and any discrepancies from the planned study have been explained.

## Data availability

The clinical codes used to define study variables are available from the corresponding author, AR, on request. Access to data is available only once approval has been obtained through the individual constituent entities controlling access to the data. The primary care data can be requested via application to the Clinical Practice Research Datalink.

## Analytic code availability

The clinical codes used to define study variables and the analytic code are available from the corresponding author, AR, on request.

